# The immune cell dynamics in the peripheral blood of cHL patients receiving anti-PD1 treatment

**DOI:** 10.1101/2024.05.15.24307370

**Authors:** Vanessa Cristaldi, Lodovico Terzi di Bergamo, Lucrezia Patruno, Marinos Kallikourdis, Giada Andrea Cassanmagnago, Francesco Corrado, Eleonora Calabretta, Adalgisa Condoluci, Martina di Trani, Daoud Rahal, Gianluca Basso, Clelia Peano, Alex Graudenzi, Marco Antoniotti, Davide Rossi, Carmelo Carlo-Stella

## Abstract

Checkpoint blockade therapy (CBT) involving anti-PD1 antibodies represents the standard approach for cHL patients who do not respond to second-line therapy. Nonetheless, only 20% of relapsed/refractory (R/R) cHL patients treated with CBT achieve complete remission. In this study, we extensively examined the immune dynamics in eight R/R cHL patients treated with CBT, consisting of four complete responders (CR) and four experiencing disease progression (PD), by single cell analysis of peripheral blood mononuclear cells (PBMCs). Our unique approach encompassed longitudinal analysis with three time points, providing a comprehensive understanding of the evolving immune responses during anti-PD1 therapy. Through gene expression profiling, we identified a stable and distinctive KLRG1+/ FOS+/JUN+/GZMA+/CD8+ T cell phenotype in patients achieving complete responses. This specific CD8+ T cell subset exhibited sustained activation, underscoring its potential pivotal role in mounting an effective immune response against cHL. Furthermore, T cell receptor (TCR) analysis revealed that in responder patients there is clonal expansion between TCR clonotypes specifically in the KLRG1+/FOS+/JUN+/GZMA+/CD8+ T cell subset. Our longitudinal study offers unique insights into the complex immune dynamics of multiply relapsed/highly pre-treated cHL patients undergoing anti-PD1 therapy.

## Introduction

Classic Hodgkin Lymphoma (cHL) is a distinct subtype of lymphoma characterized by the presence of Hodgkin and Reed-Sternberg (HRS) cells within a background of inflammatory infiltrate^1,2^. The immune microenvironment plays a critical role in the pathogenesis and progression of cHL^3,4^. HRS cells evade immune surveillance through various mechanisms, including the expression of programmed death-ligand 1 (PD-L1) and secretion of immunosuppressive cytokines^5^.

The tumor microenvironment of cHL is characterized by a diverse array of immune cells, including T cells, B cells, macrophages, eosinophils, and mast cells^6^. CD4+ T helper cells are abundant within cHL tumors^7,8^, but they often exhibit an exhausted phenotype marked by high expression of immune checkpoint receptors, such as programmed cell death protein 1 (PD-1)^9^. Regulatory T cells (Tregs) are also enriched within cHL lesions, contributing to immunosuppression and tumor immune evasion^10,11^.

Immunotherapy has revolutionized the treatment landscape of cHL, particularly through the use of immune checkpoint inhibitors targeting the PD-1/PD-L1 axis^12,13^. Pembrolizumab and nivolumab, anti-PD-1 antibodies, have demonstrated remarkable efficacy in relapsed or refractory cHL, leading to durable responses in a significant proportion of patients^14–15^.

The rationale for targeting PD-1/PD-L1 signaling in cHL stems from the overexpression of PD-L1 on HRS cells and the presence of PD-1-expressing exhausted T cells within the tumor microenvironment. By blocking the PD-1/PD-L1 interaction, immune checkpoint inhibitors unleash the cytotoxic activity of T cells against cHL cells, leading to tumor regression.^16^

More recently, clinical trials have demonstrated the efficacy of immune checkpoint inhibitors as monotherapy or in combination with chemotherapy in additional settings^17^, including first relapsed/refractory disease^18^, frontline treatment and as maintenance therapy following transplantation^19,20^. However, not all patients respond to immunotherapy, highlighting the need for accessible biomarkers to identify responders and strategies to overcome resistance mechanisms.

Our research aims to investigate whether alterations in the peripheral blood T cell pool can serve as a biomarker for predicting response to immunotherapy in patients with R/R cHL. Through single-cell RNA sequencing (scRNA-seq)^21^ and single cell TCR sequencing (scTCR-seq)^22^, we obtained high-resolution transcriptomic profiles of peripheral blood mononuclear cells of patients with R/R cHL, deconvoluted the T cell subtypes and clonotypes and correlated them with response to immunotherapy.

## Materials and methods

### Patients Recruitment

The study was retrospective in nature and included a total of eight patients with R/R cHL, aged >18 years, who were enrolled in the CheckMate 205 trial ^23^and treated with Nivolumab (Table 1). Staging and disease response were assessed according to the Lugano 2014 criteria^24^. The study was conducted in accordance with International Conference on Harmonization for Good Clinical Practice guidelines and the Declaration of Helsinki^25^. Written informed consent was obtained from all patients before enrollment. Patients were administered Nivolumab at a dose of 3 mg per kilogram of body weight^26^ every 2 weeks. All patients received treatment at Humanitas Research Hospital.

**Table 1.**
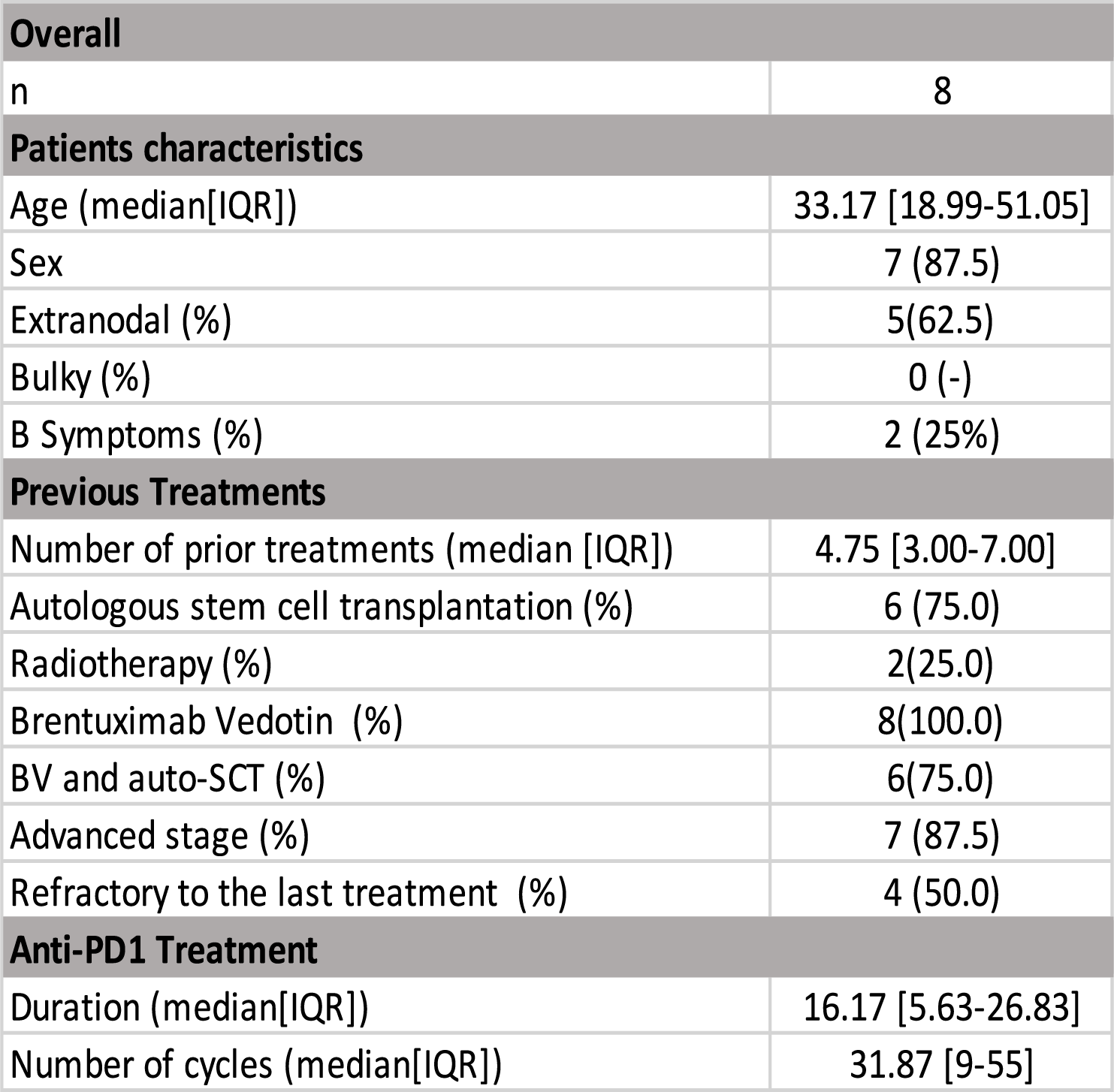
Pa*ent Feature Overview Table.

For each patient, we collected three samples of PBMCs: before cycle 1 (Baseline or Pre-Treatment stage), before cycle 5 (Intermediate stage) and before cycle 10 (Post Treatment stage) with anti-PD1 immunotherapy (except for one PD patient whose cycle 5 we do not have) (Fig.1a). These patients were categorized into two response groups based on their response at cycle 10: four patients who remained in progressive disease (PD) and four patients who achieved a complete response (CR).

**Figure 1.**
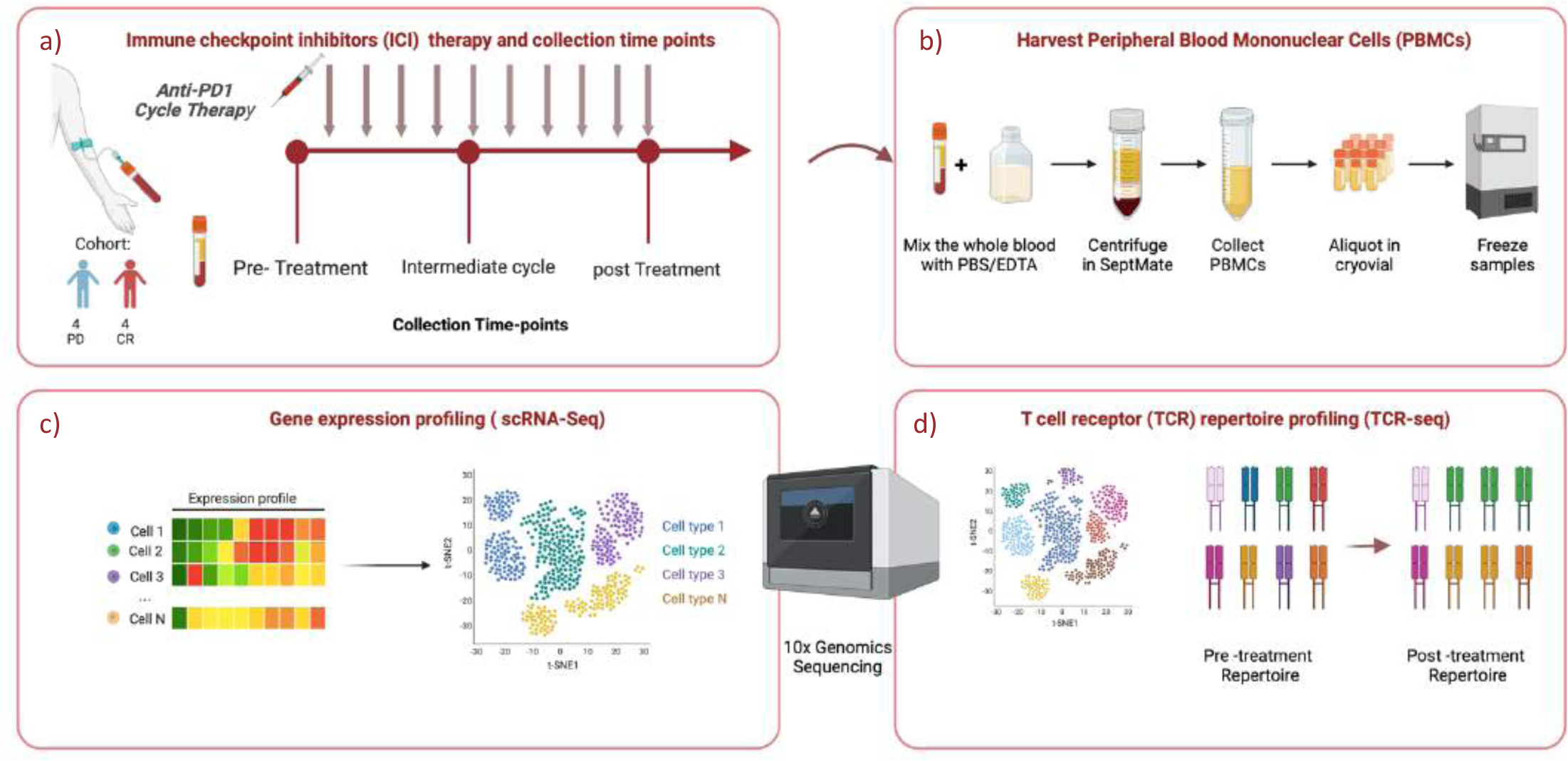
Overall study design. **a-b)** Eight patients were included in the study, from whom peripheral blood were collected at three different time points: before treatment initiation (Pre-treatment), after the 4th cycle of therapy (Post 4 cycles), and following the 8th or 9th cycles of treatment (Post 8-9 cycles). Four exhibited progression disease (PD), and four achieved a complete response (CR). **c-d)** PBMCs were assayed by single-cell RNA sequencing and T-cell receptor sequencing (TCR-SEQ).

### Library Preparation and single-cell RNA sequencing

In total an average of 5000 cells per sample were loaded into a Chromium Single Cell 3’ Chip kit v2 (PN-120236) and processed according to the Chromium Single Cell 3’ Reagent kit v2 User Guide. Libraries were constructed using the Single Cell 3’ Library and Gel Bead Kit v2 (PN-120237) and Chromium i7 Multiplex Kit (PN-120262). All single cell libraries were pooled and sequenced on Illumina NextSeq550. CellRanger software (v2.1.0; 10X Genomics) was used to demultiplex the raw data, generate quality metrics, and generate per-gene count data for each cell. Count data from 8 r/r cHL samples were pooled together, including only features with expression data found in at least 3 cells, and including cells with at least 200 features observed, cells were filtered out if they had >20% reads aligning to mitochondrial genes, or if they had >2500 features detected. Exclusion of run specific effects was observed using UMAP plots. Clustering was performed using the FindClusters function from the Seurat^27^ package in R, and the clustree package was used for determining the best number of clusters. The total list of genes used for clusters annotation was composed of previously described genes signatures^18^ and a list of manually curated markers from the literature. Unsupervised hierarchical clustering was performed on this set of markers, and clusters were manually annotated based on their gene expression levels. Differential expression between patients and between class of response were performed in selected cell populations using the seurat package.

### T Cell Receptor sequencing

The same samples analyzed with scRNA-seq, were also analyzed for scTCR-sequencing. 10X Genomics standard protocol was applied and the reagents for the Chromium Single-cell 5’ Library and V(D)J library (v2.0 Chemistry) were used. Using a microfluidic technology, single-cells were captured and lysed, mRNA was reverse transcribed to barcoded cDNA using the provided reagents (10X Genomics). 14 PCR cycles were used to amplify cDNA. Part of the material was target-enriched for TCRs and V(D)J library was obtained according to manufacturer protocol (10X Genomics). Barcoded VDJ libraries were pooled and sequenced by an Illumina NextSeq 550 Sequencer. Single-cell TCR sequencing data were processed by the Cell Ranger software pipeline (v2.1.0; 10X Genomics). The TCR sequence data were processed using Scirpy^28^ (Single-Cell Immune Receptor Profiling), employed for data preprocessing and advanced analysis of single-cell T-cell receptor repertoires. Key functionalities of Scirpy encompass raw data demultiplexing, assignment of T-cell receptor variable region sequences, generation of clonal frequency matrices, and analysis of clonotype distributions. Specifically, Scirpy was utilized to identify and quantify clonal diversity within T-cell populations, ascertain the presence of dominant clones, calculate diversity metrics, and assess repertoire stability over time.

### Statistical Analysis

Statistical analyses were conducted using GraphPad Prism (version 9) in conjunction with R version 4.0.3. Specifically, for group comparisons, the t-test and Mann-Whitney test were employed, depending on the data distribution and specific analysis conditions.

## Results

### Patients’ characteristics and response to therapy

Patients (n=8) had a median age of 33 years (interquartile range 19-51), and 7 (87.5%) were males. Before treatment, 87.5% of patients (n=7) showed an advanced disease (Stage IIB-IV), with extranodal involvement present in 5 (62%) cases. Notably, the study group consisted of highly pre-treated patients, with a median of 5 previous lines of therapy (**Table1**). Six (75%) patients had previously received autologous stem-cell transplantation and all had received Brentuximab-Vedotin. Patients were treated with Nivolumab for a median of 16 cycles (interquartile range 6-27) and classified as responsive or unresponsive to the PD1 blockade based on the best overall response (BOR) and the duration of response (DOR). The best overall response was defined as the best response between the first dose and progression.^29^ Patients who achieved a best complete response were considered responsive (n=4) while patients with primary progressive disease (n=1) or best partial response lasting less than 6 months (n=3) were considered unresponsive. Among patients achieving CR, 3 (75%) underwent consolidative allogeneic transplantation (allo-SCT). The remaining patient exhibited a prolonged DOR (15.5 months). Patients achieving a best partial response (n=3) showed a limited median DOR of 4 months. No gross difference in the distribution of main clinical features was observed between the two groups (**Supp Table 1**).

### Immune cells dynamics during Nivolumab therapy

With limitations imposed by sample size, after performing an integrated analysis with all samples, we compared pre-treatment, during, and post-treatment PB cell types between responsive and non-responsive patients (Fig.2a), observing a trend in which populations of lymphocytes such as naïve CD4+ T cells (CD4+TN), CD4+ central memory (CD4+CM), CD8+ T cells, γδT cells and B cells are enriched in responding patients, in all phases of treatment, suggesting a synergistic response of these cells against tumor cells, but no statistical significance was observed. On the contrary, populations such as CD14+ and CD14- and CD16+ monocytes are more abundant in non-responders during therapy and already at pre-treatment stage. (Fig.2b).

**Figure 2.**
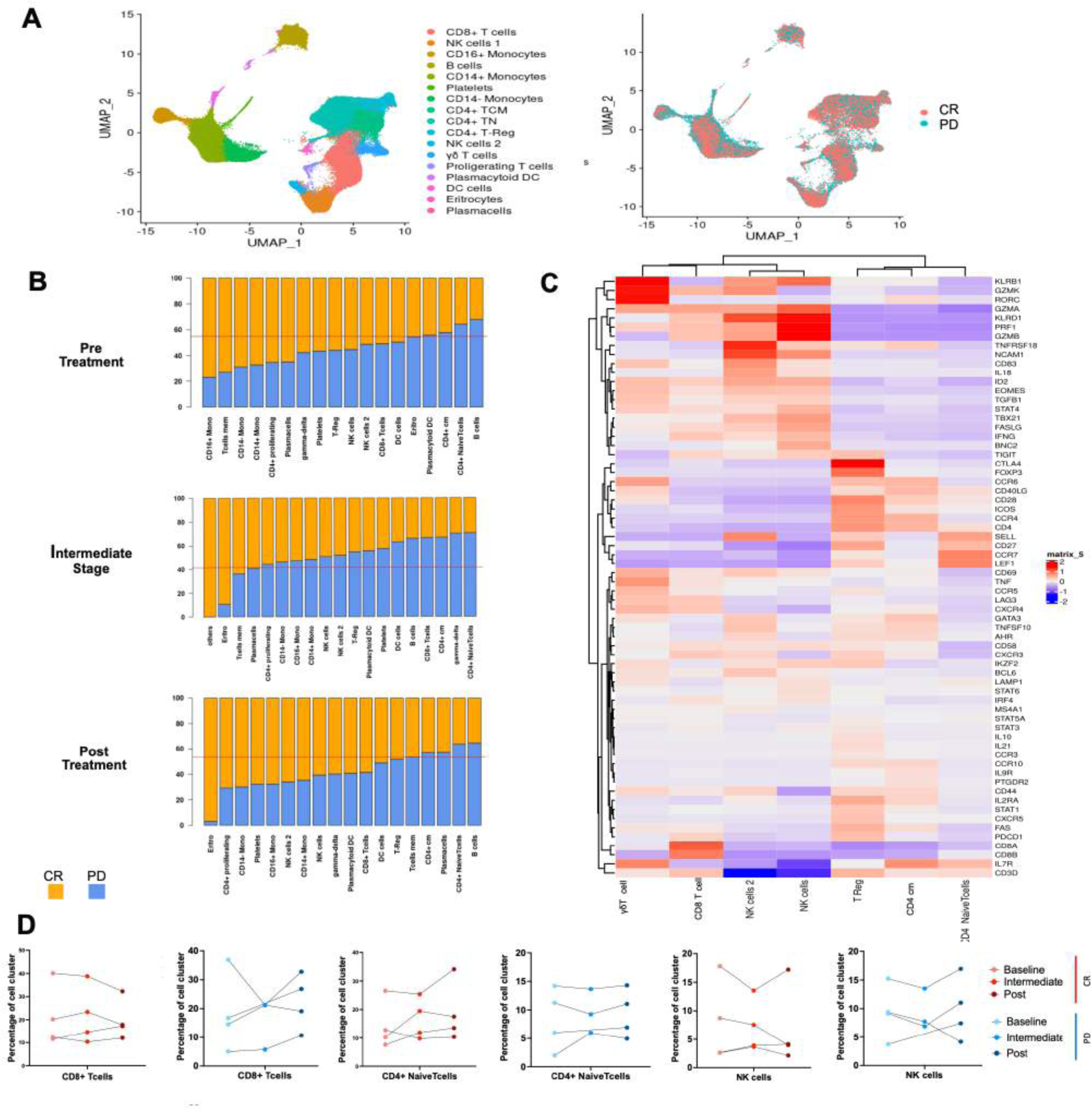
Distinct patterns of gene expression between PD and CR patients under anti-PD1 therapy a) Two-dimensional similarity map (UMAP projection) of single-cell gene expression of XXX cells from 8 r/r cHL. Cells are colored according to the PhenoGraph cluster (left) and by class of final response to treatment (right) **b)** Barplot with proportion of each cell cluster, according to time-point for both the PD and CR class. The horizontal red line represents the expected number of cells based if the clusters were equally represented in CR and PD cHL **c)** Heatmap summarizing the mean expression (normalized and log-transformed) of selected canonical markers for each cluster (γδ Tcell, gammadelta T cells; CD8T cells; NK cells 2, Natural Killer cells type 2; NK cells, Natural Killer cells; T-Reg, regulatory T cells; CD4 CM, central memory T cells; CD4 TN, naïve T cells). **d)** Percentage of cells in each cluster during time (Pre-Treatment, Intermediate stage and Post-Treatment) in both classes of patients.

Furthermore, following the application of signatures associated with the type I interferon response to inflammation on patients’ peripheral blood mononuclear cells (PBMCs), a noteworthy observation emerged. Specifically, in PD patients, the monocytic subgroup demonstrated a remarkably distinct module score linked to the pathway associated with response to type I interferon stimulation during the pre-treatment phase (GO: 0034340 from human GSEA dataset) (Supplementary Figure 4)

Focusing our attention on CD8+ T cells, we conducted differential expression gene (DEG) analysis across all peripheral blood mononuclear cell (PBMC) populations. (Supplementary Figure 3a-b-c). Over time, CD8+ T cells from responsive patients (CR) exhibited a significant (cut-off for log2FC >2; P=10e-32) upregulation of effector/activation genes such as *KLRG1*, *FOS*, *JUN*, and *GZMA* (Fig.3). Co-expression of *KLRG1*,*FOS*, *JUN*, and *GZMA* remained constant during all time-points in responsive patients. Using genes identified from differential gene expression analyzes between CD8+ cells from treatment-responsive and non-responsive patients, a Gene Set Enrichment Analysis (GSEA) was conducted to identify the gene pathways involved. Complete response (CR) patients showed significant enrichment (p value adjust < 0.05) of gene sets related to T cell receptor (TCR) activation, cellular activation and T cell activation, which was also confirmed after treatment. At midcycle, CR patients showed significant up-regulation of processes related to T cell activity, including the assembly and activity of Major Histocompatibility Complex (MHC) class I.

**Figure 3:**
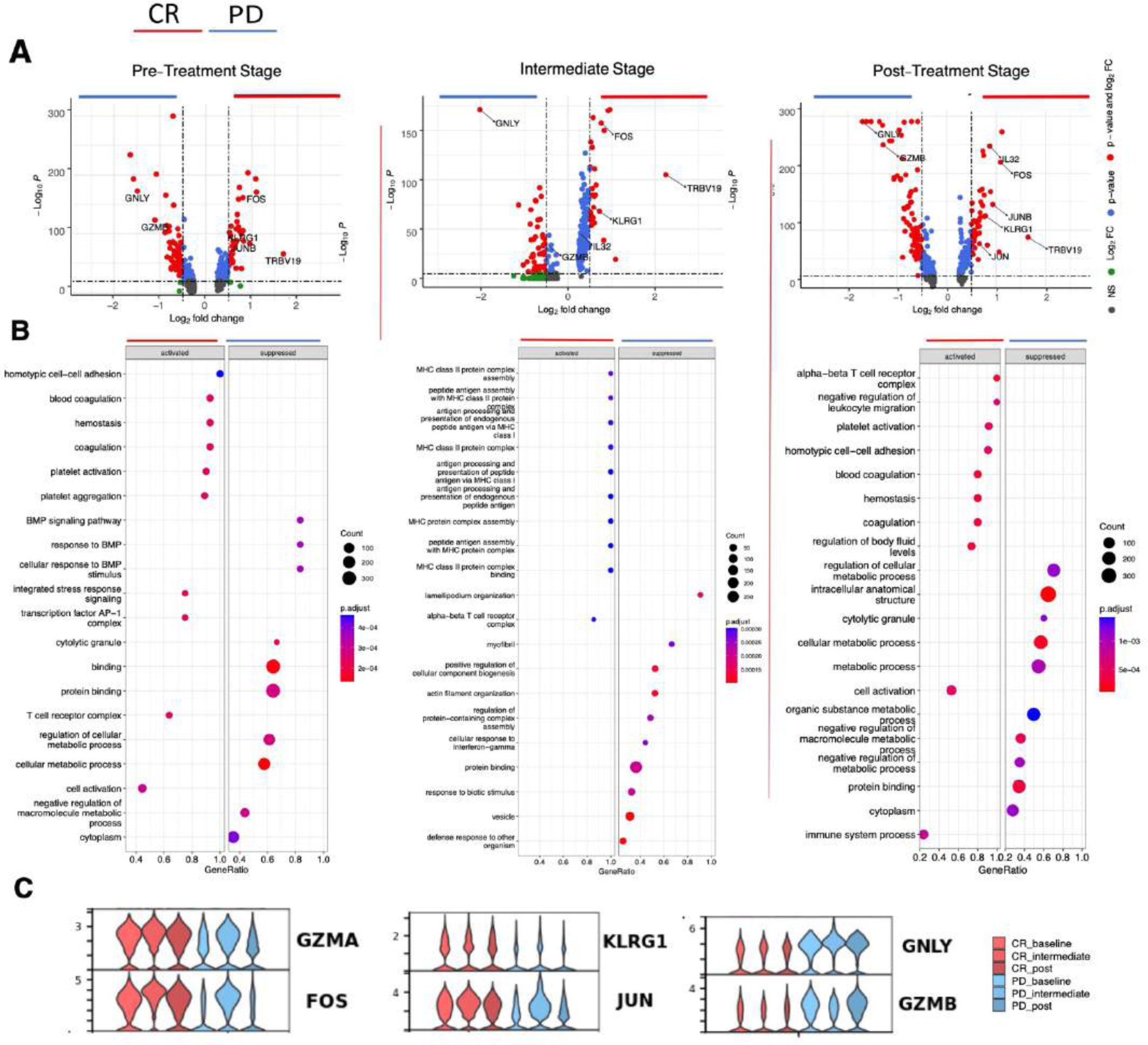
CD8+ T cells activation in CR patients a) Volcano plot showing differentially expressed genes between CD8+ T cells of responsive patients (CR) vs non responsive patients (PD) in pre-treatment (left), intermediate (middle) and post-treatment (right). Significant genes are labeled in red (P value <0.05 and absolute log2 fold change ≥0.5). Selected important genes for T cells are labelled. **b)** Gene Set Enrichment Analysis (GSEA) of differentially expressed genes between CR and PD patients for each time-point. Dots are coloured based on significance, and their size is proportional to the number of genes belonging to each gene set **c)** Violin plot showing the expression of KLRG1, JUN, GZMA, FOS, GNLY and GZMB in each group of patients during time.

### TCR clonal expansion observed in KLRG1+ FOS+ JUN+ GZMA+ CD8+ T Subclusters of responsive patient’s class

We conducted immunophenotypic analysis of lymphocytes through T cell receptor repertoire sequencing (TCR-seq) (10x Genomics) to better understand the immune response in patients. TCR analysis revealed a clonal expansion phenomenon, i.e. the presence of a TCR structure shared by more than 3 cells, which was found to be predominant in the CD8+ T cell cluster in both response groups (Fig.4a). Furthermore, we observed greater clonal expansion in central memory (cm) CD4+ T cells in non-responsive patients, especially in the intermediate stage. Naïve CD4+ T cells, however, mainly showed associated expansion of individual clones during treatment and no expansion of clones overall over time (Fig.4b). A more detailed analysis of TCR structures revealed an overlap of the expanded CD8+ TCR clonotypes in responsive patients with the CD8+ T cell subcluster characterized by KLRG1+ and GZMA+ phenotype, previously identified in scRNA-seq analysis. This overlap suggests a correlation between the clonal expansion of CD8+ TCRs and the T cell phenotype associated with treatment response (Fig.4c).

**Figure 4:**
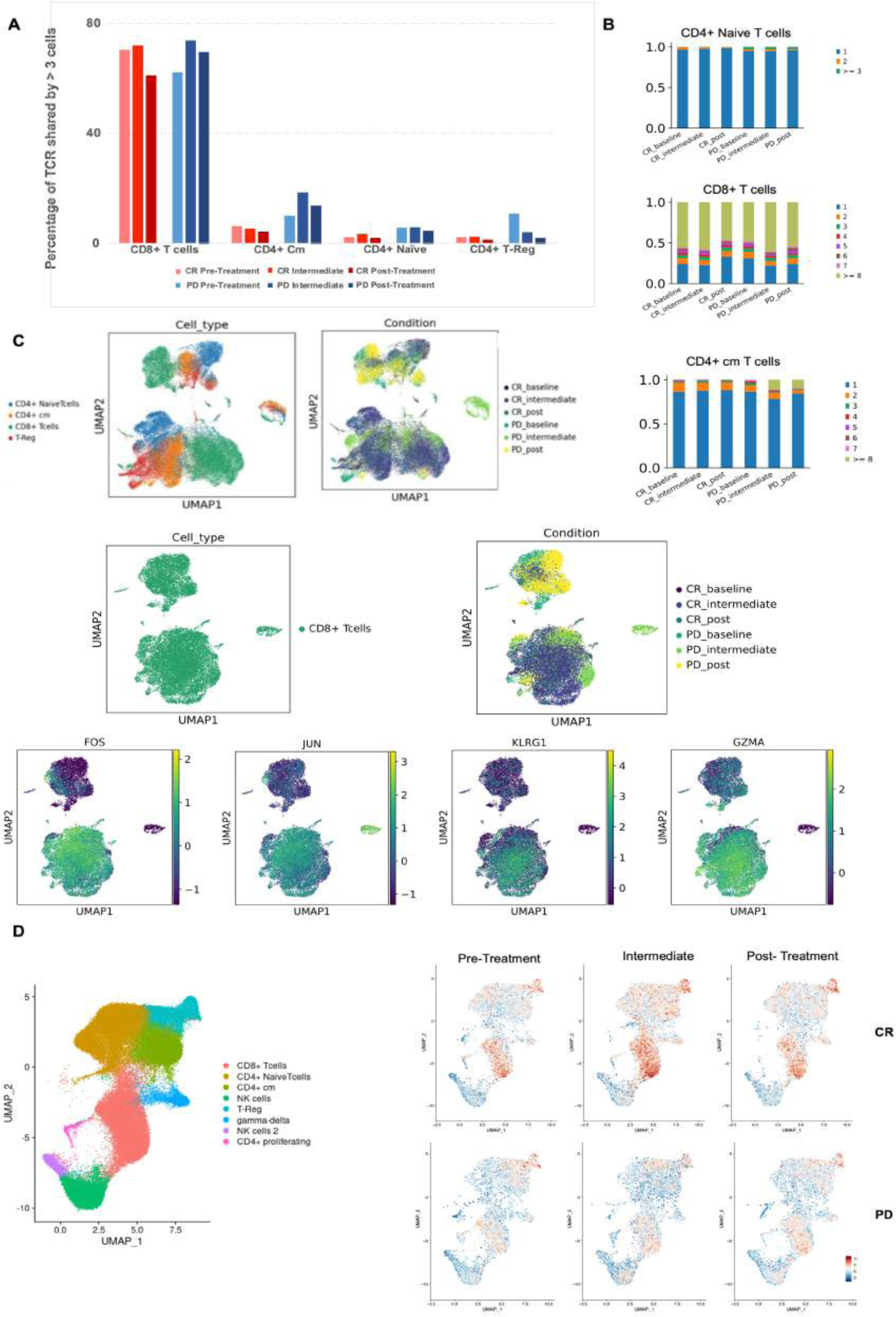
TCR clonotype tracking and clonotype expansion during time a) Barplot showing the percentage of T cell receptor (TCR) shared by more than 3 cells in each cluster, comparing CR and PD. **b)** Detailed TCR clonotype expansion from 0 to 1.0 (100%) for CD4 central memory, CD8 T cells and Naïve T cells, in all patient conditions. **c)** UMAP showing TCR clusters based on their CD3 region sequence, colored by cell type, and **d)** by groups of patients during time. **e)** UMAP showing T cells with TCR structure annotated and overlapped markers genes. **f)** UMAP and signature related to TCR activation (GOCC: Alpha-beta TCR-Complex-GSEA dataset).

To further evaluate this correlation, we applied a TCR activation signature (GOCC: “Alpha Beta T cell receptoc complex” from human GSEA dataset), which includes genes associated with the alpha-beta T cell receptor complex, to the dataset (Fig.4d). The results show that the module score is higher in CD8+ TCRs from responding patients, indicating an association between clonal expansion of CD8+ TCRs and treatment response in patients with complete response (CR).

## Discussion

In this genetic and immunophenotypic profiling study of PBMC populations we investigated the immune dynamics of patients with relapsed/refractory classical Hodgkin lymphoma (r/r cHL). It is essential to recognize that these patients, despite having demonstrated a non-response to previous treatments, were still subjected to various therapies before anti-PD1 treatment (Table 1). Of relevance, our cohort included heavily pre-treated patients, who received a high number of therapies prior to PD-1 blockade. Given these considerations, our study observed the KLRG1+, GZMA+ phenotype in CD8+ T cells in patients who achieved a complete response (CR) to anti-PD1 therapy.

These findings suggest that this specific subset of CD8+ T cells could play a fundamental role in predisposing the patient to a better response to anti-PD1 treatment. Furthermore, it is interesting to note that this phenotype remained stable throughout time, suggesting consistent and robust activity of CD8+ T cells with this specific phenotype.

These genes are known to play a role in the activation of CD8+ T cells, enhancing their antitumor function^30^. *KLRG1*, Killer Cell Lectin-Like Receptor G1, is predominantly expressed by CD8+ effector T cells and influences immunological memory^31,32^. The genes FOS and JUN indicate an activation and differentiation of CD8+ T cells towards a more effector state^33^ and GZMA is involved in cytotoxic mechanisms^34^.

The significant Increase in the expression of genes such as GZMB (granzyme B) and GNLY (granulysin) in CD8+ T cells of patients unresponsive to anti-PD1 treatment could reflect an attempt by the immune system to fight the tumor through the activation of CD8+ T cells. However, despite the increase in these genes, the lack of response may suggest that other factors, such as the immunosuppressive tumor microenvironment or the presence of immune evasion mechanisms, may limit the effectiveness of the cytotoxic activity of CD8+ T cells.

Furthermore, the expression of GZMB and GNLY could indicate a state of chronic activation of CD8+ T cells^35^, which could lead to exhaustion and dysfunction of T cells, hindering their effective role in eliminating tumor cells^36^. Therefore, these findings highlight the importance of a deeper understanding of the mechanisms involved in Hodgkin lymphoma and the response to immunotherapies to develop more effective strategies for the treatment of non-responsive patients.

Regarding the TCR analysis, it is interesting to note that we did not observe significant differences in clonal expansion between CD8+ T cells from CR and PD patients. However, the most significant aspect of this TCR data lies in the precise overlap between TCR clonotypes exhibiting clonal expansion (>3 cells) in CR patients and the KLRG1+, FOS+, JUN+, GZMA+ CD8+ T cell subset. This detail is crucial as it suggests that this specific subset of CD8+ T cells could be relevant in mediating the immune response in patients who respond to anti-PD1.

This alignment strengthens the robustness of our findings by demonstrating a consistent relationship between TCR specificity and cellular characteristics. Further research is needed to fully elucidate the functional significance of these TCR phenotypic associations and to determine how they can be exploited for therapeutic benefit.

Overall, this work provides further evidence of the dynamic and complex nature of the immune response induced by anti-PD1 therapy. The expansion of specific CD8+ T cell clones, especially within responding patients, suggests their potential role in driving an effective anti-tumor response. However, the simultaneous clonal expansion of additional CD8+ T cell clones in all patients indicates a more complex interaction of immune cell populations during treatment.

Further investigation into the functional properties of these expanded T cell clones and their interactions with other immune cells would be essential to fully understand their contribution to treatment outcomes and guide the development of personalized immunotherapeutic strategies for cancer patients undergoing anti-PD1 therapy.

## Data Availability

All data produced in the present work are contained in the manuscript.

## ACKNOWLEDGMENTS

This work was supported in part by a grant from the Italian Association for Cancer Research (AIRC, grant #20575 to CC-S). Vanessa Cristaldi was supported by Post-doctoral Fellowships 2023 by Fondazione Umberto Veronesi.

## AUTHORSHIP CONTRIBUTIONS

Vanessa Cristaldi conceived, performed, analyzed the experiments and wrote the manuscript. Carmelo Carlo-Stella has acquired financing. Davide Rossi and Carmelo Carlo-Stella conceived and revisioned the project. Vanessa Cristaldi, Lodovico Terzi di Bergamo, Lucrezia Patruno, Alex Graudenzi and Marco Antoniotti contributed to the bioinformatic analysis. Francesco Corrado, Daoud Rahal and Eleonora Calabretta collected and reviewed clinical data. Gianluca Basso and Clelia Peano provided support in the sequencing analyses. All others provided critical input and revised the manuscript.

## Figures and Tables

**Supplementary Figures 1a:**
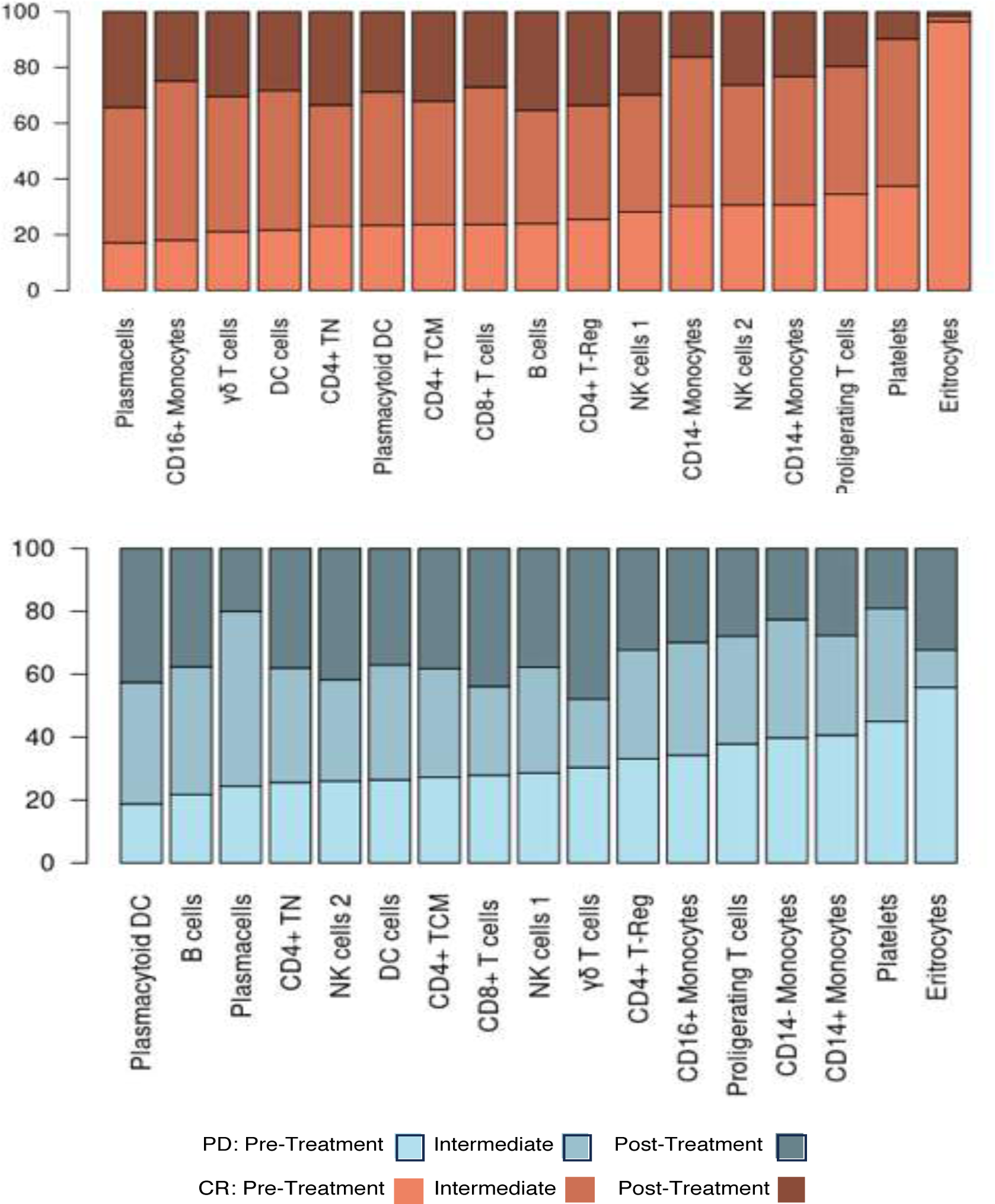
Barplot with proportion of each cell cluster in PD patients vs CR patients, in each stage of treatment. Red line represents the expected proportion if the cluster is equally represented in CR and PD samples

**Supplementary Figures 1b:**
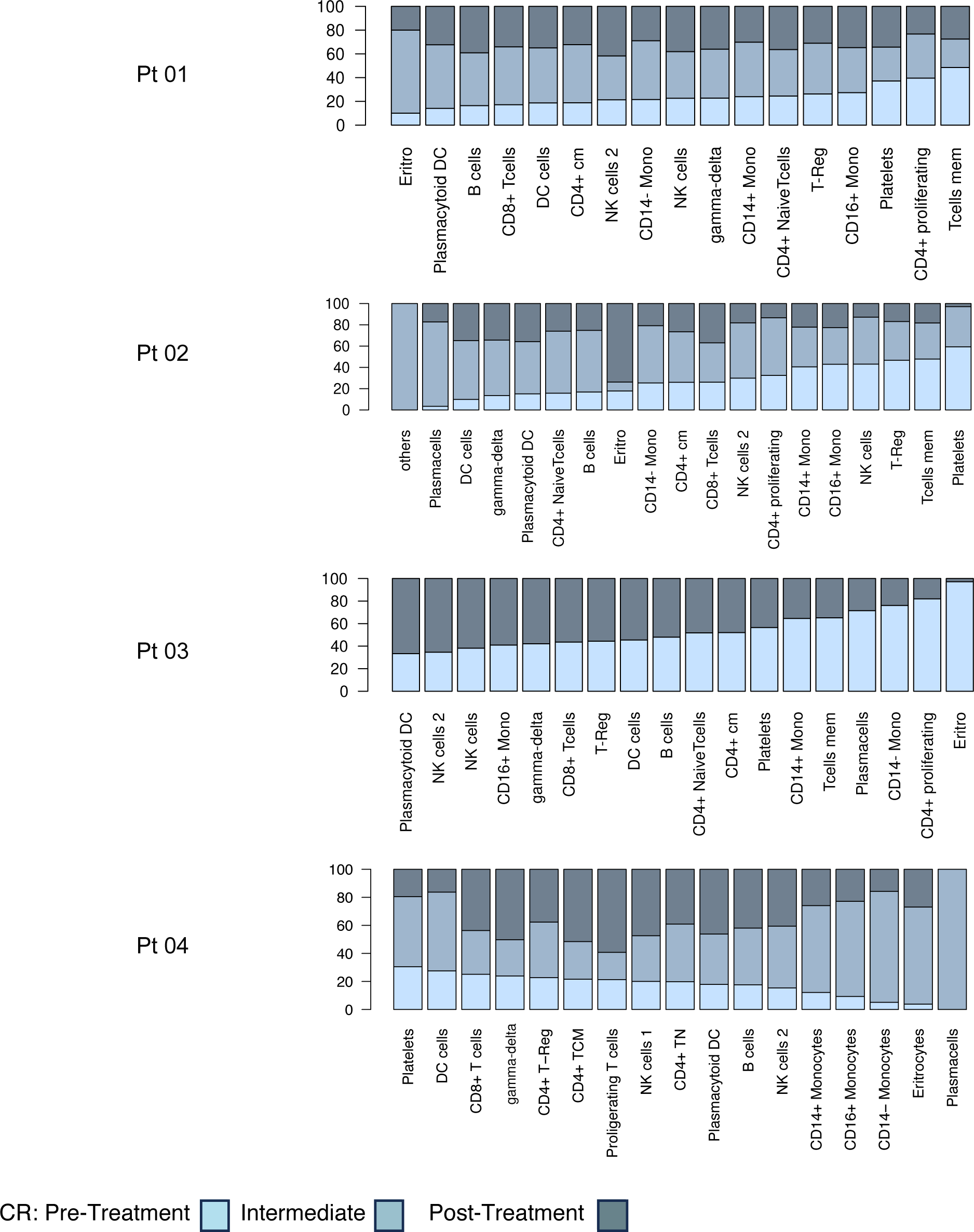
Barplot with proportion of each cell cluster in every PD patient, during each stage of treatment.

**Supplementary Figures 1c:**
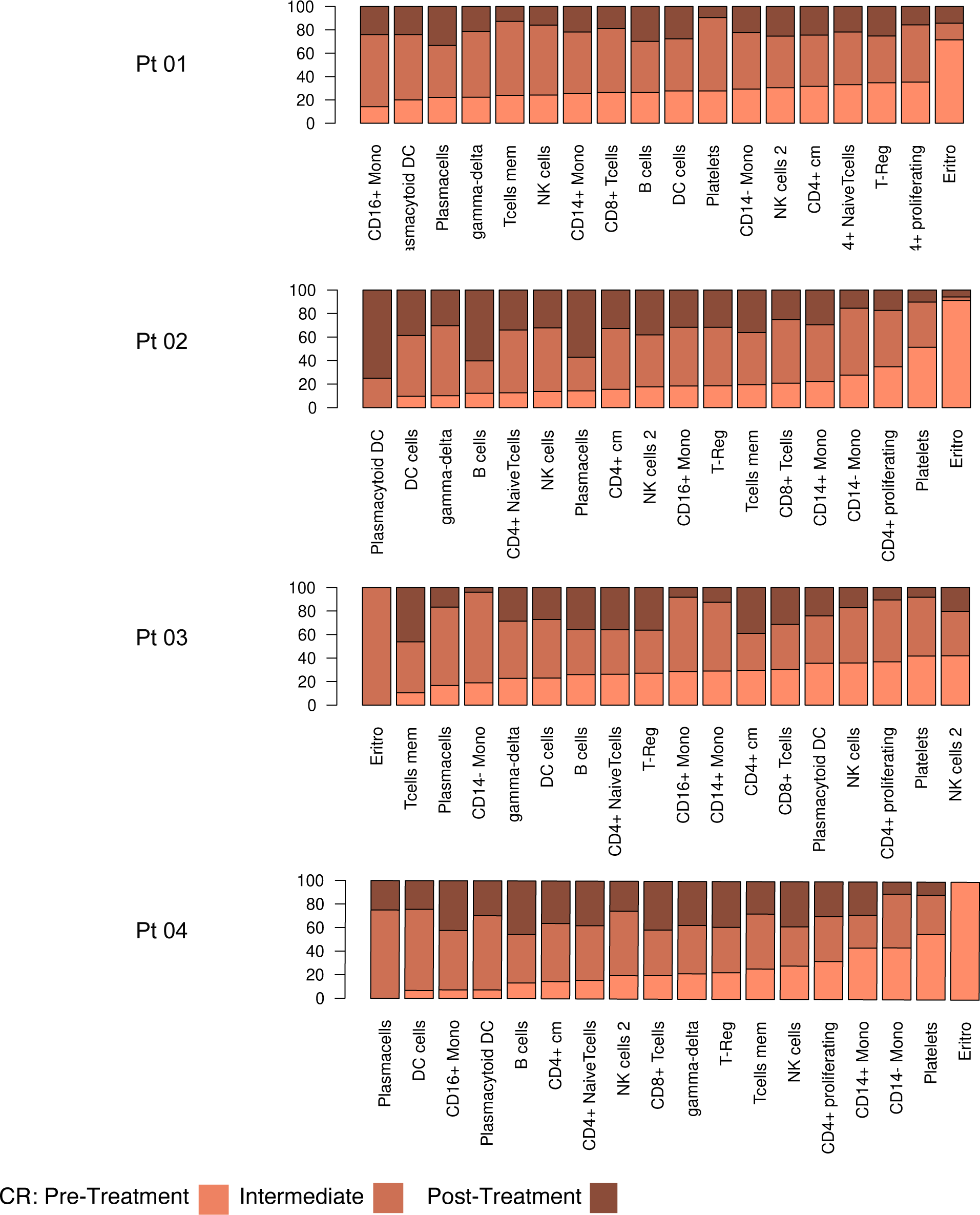
Barplot with proportion of each cell cluster in every CR patient, during each stage of treatment.

**Supplementary Figures 2:**
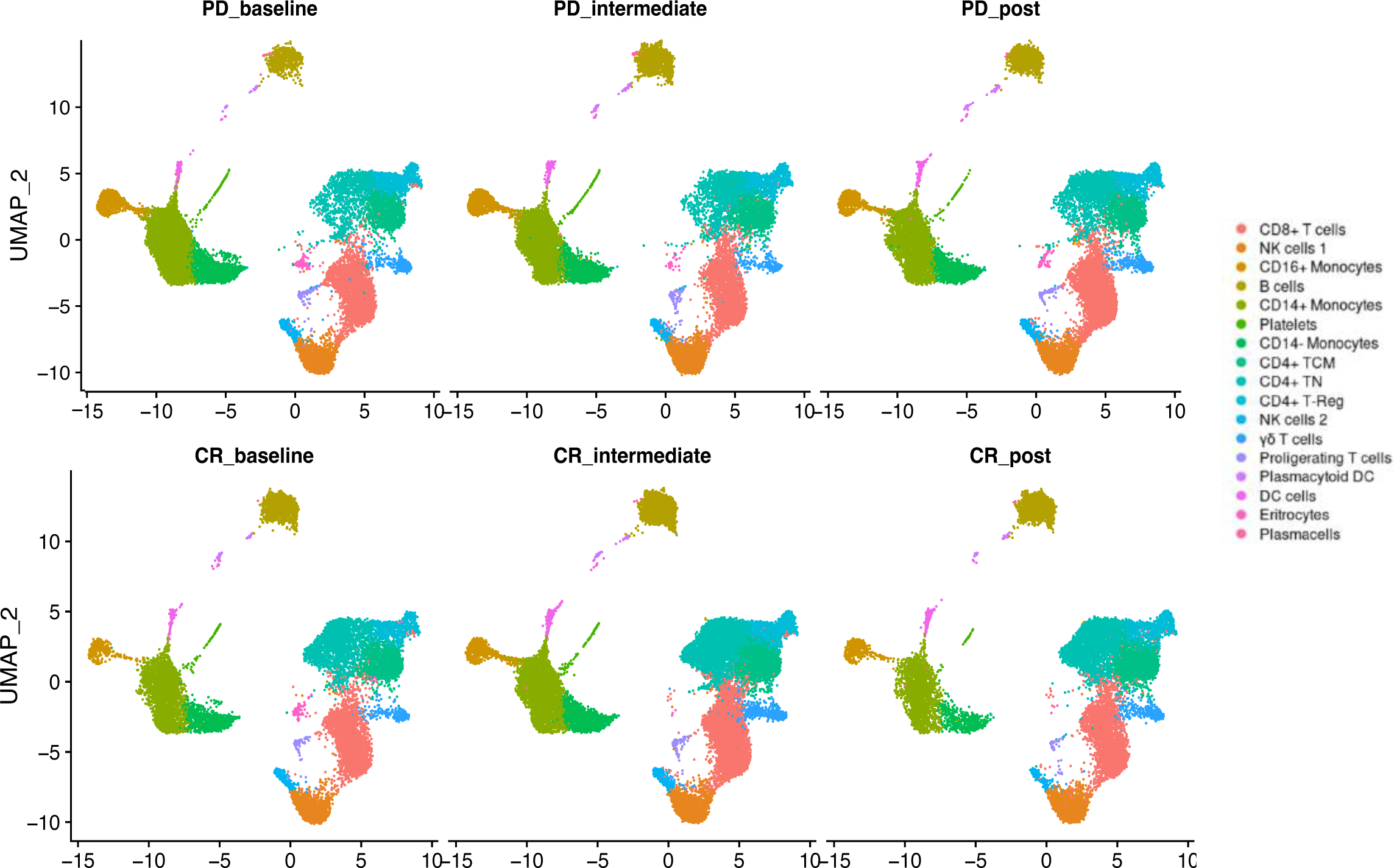
Two-dimensional similarity map (UMAP projection) of single-cell gene expression in different timepoints and different conditions. Cells are colored according to the PhenoGraph cluster. Subsets of cells are nominated according to their profile of gene markers.

**Supplementary Figures 3a:**
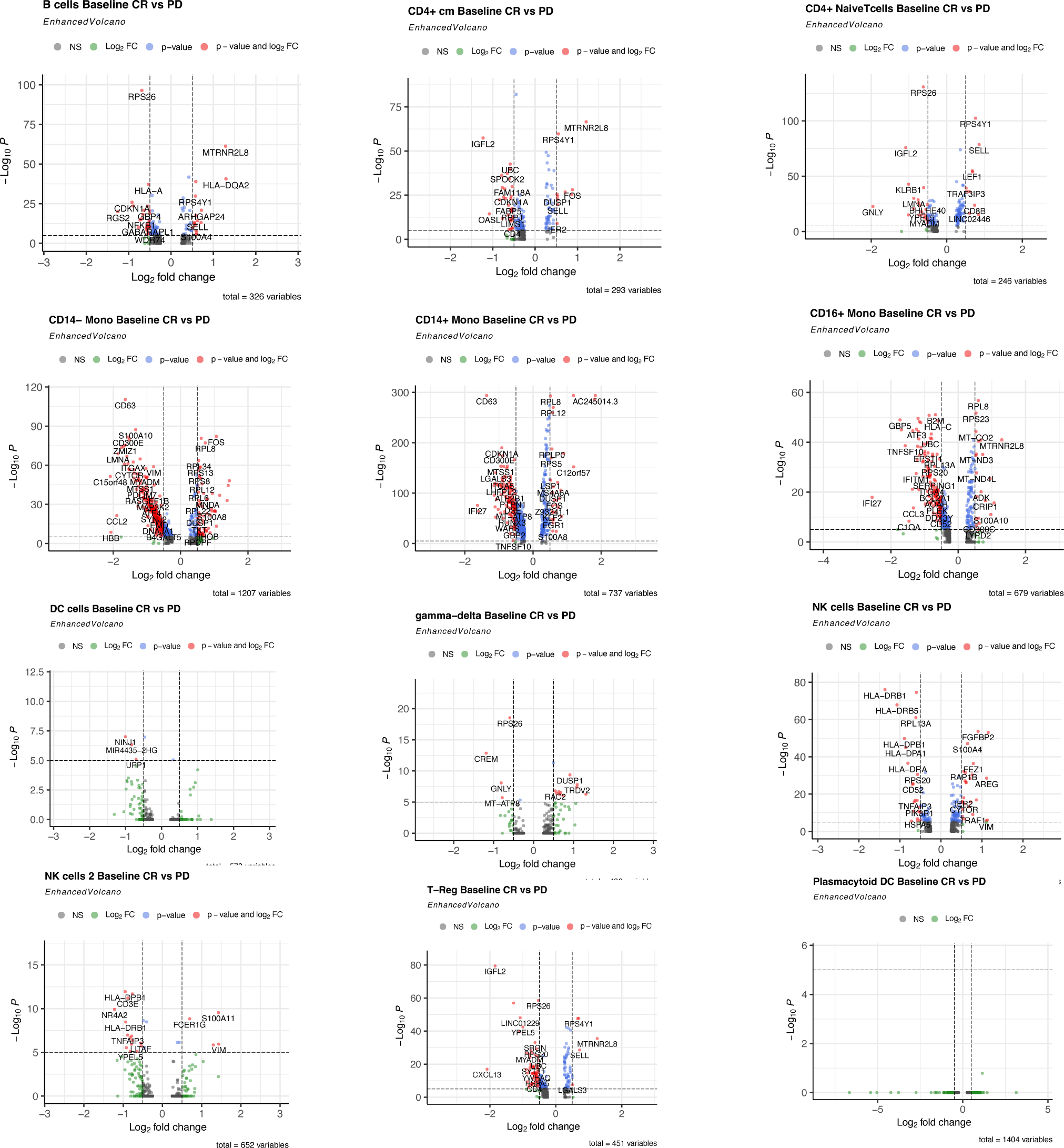
Differential expression analysis in CD8+ T cells of CR patients at pre-treatment. Volcano plot showing differentially expressed genes between each cluster cells of responsive patients (CR) vs non responsive patients (PD). Significant genes are labeled in red (P value <0.05 and absolute log2 fold change ≥0.5).

**Supplementary Figures 3b:**
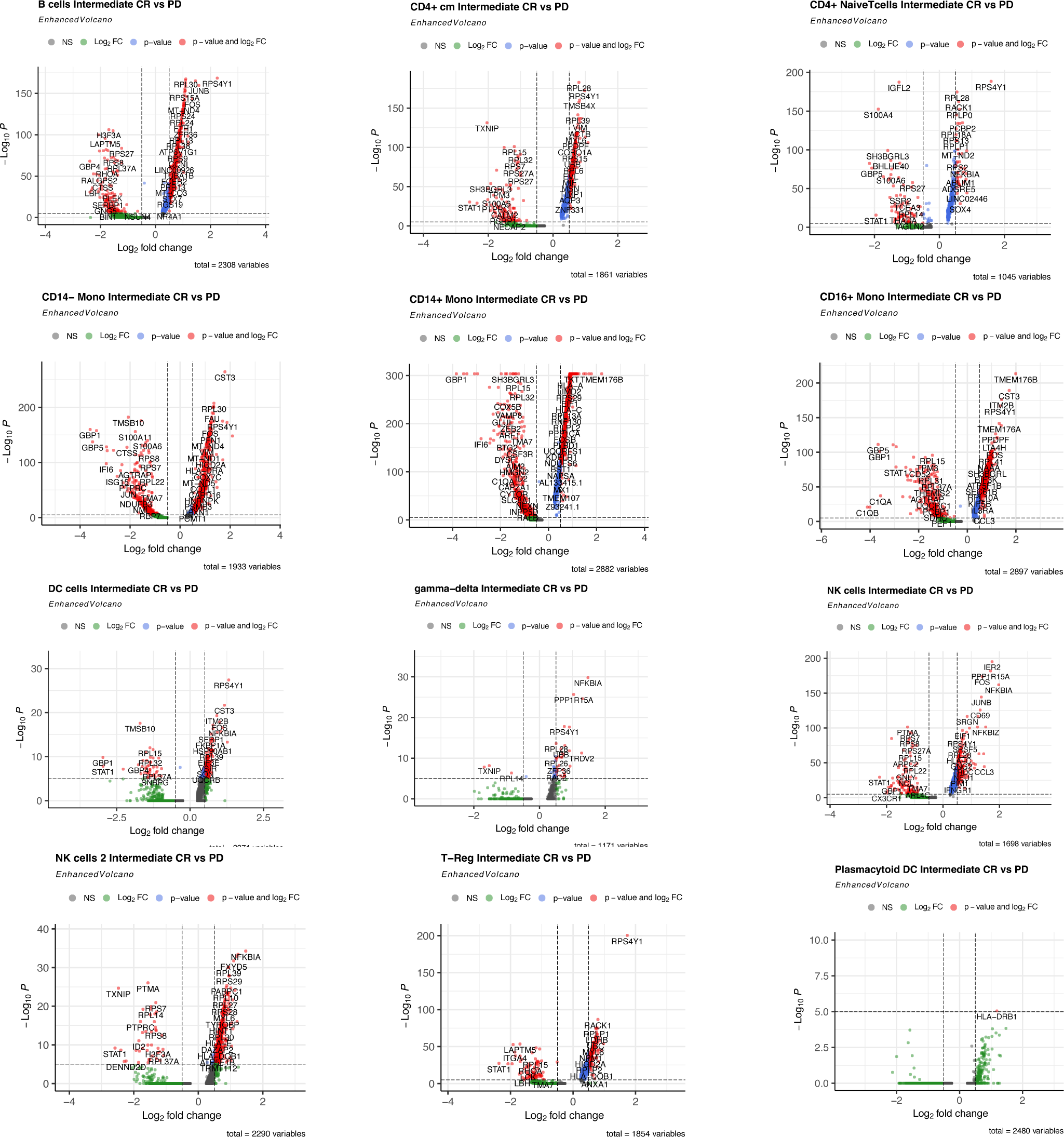
Differential expression analysis in CD8+ T cells of CR patients At intermediate stage. Volcano plot showing differentially expressed genes between each cluster cells of responsive patients (CR) vs non responsive patients (PD). Significant genes are labeled in red (P value <0.05 and absolute log2 fold change ≥0.5)

**Supplementary Figures 3c:**
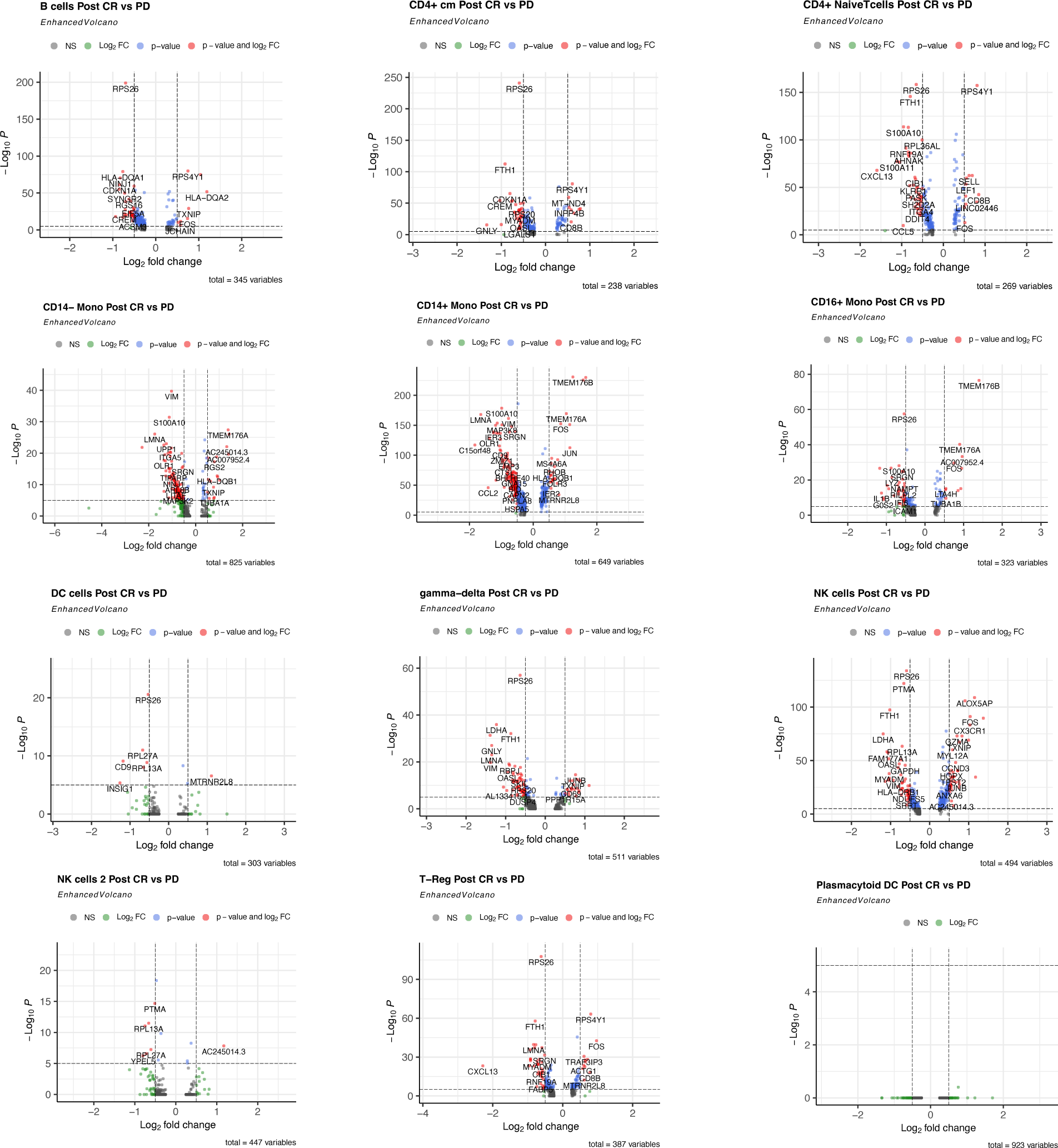
Differential expression analysis in CD8+ T cells of CR patients at post-treatment. Volcano plot showing differentially expressed genes between each cluster cells of responsive patients (CR) vs non responsive patients (PD). Significant genes are labeled in red (P value <0.05 and absolute log2 fold change ≥0.5).

**Supplementary Figure 4:**
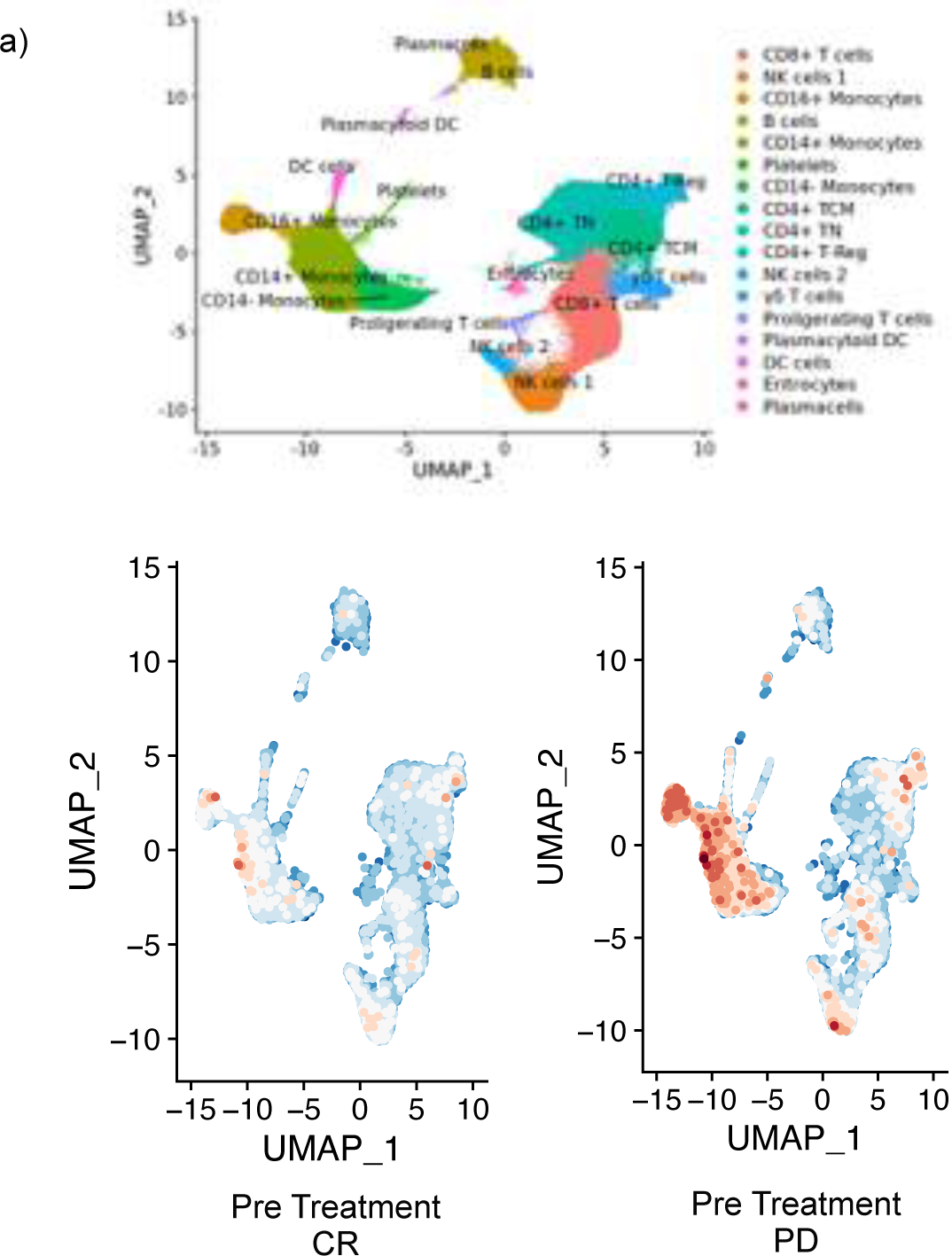
**a)** Combined two-dimensional similarity map (UMAP projection) of single-cell gene expression for all samples in the study cohort. **b)** UMAP projections with cells colored by module score analysis of pathway linked to the response to type I interferon at pre-treatment stage (GO:0034340), comparing CR baseline stage and PD Baseline stage.

**Supplementary table 1.**
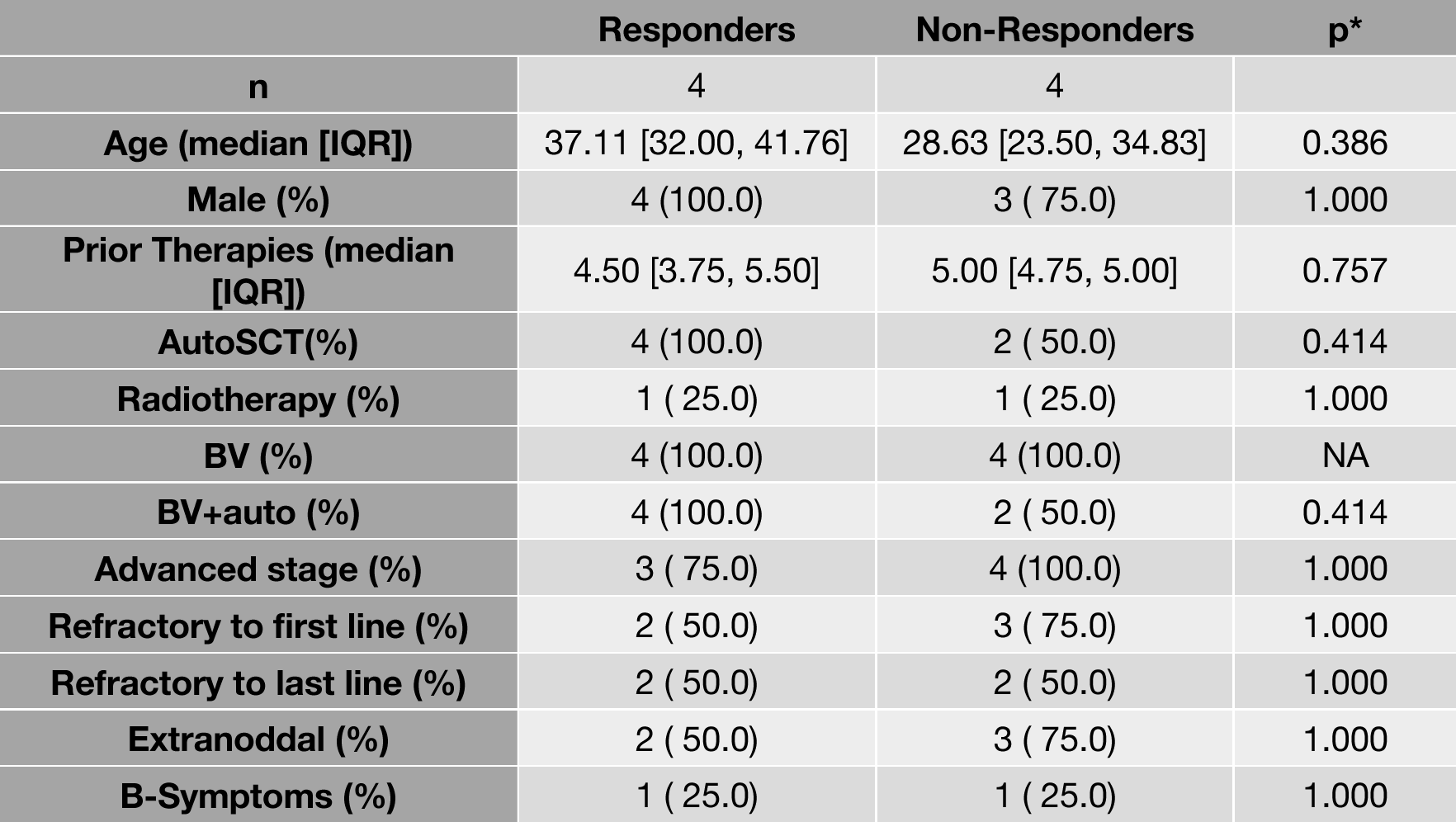
Table of main clinical covariates between the two groups of patients. p*= Categorical variables are assessed using the Fisher’s test (p-value), while continuous variables undergo analysis with the Wilcoxon-Mann-Whitney test.

|  | **Responders** | **Non-Responders** | **p\*** |
| --- | --- | --- | --- |
| **n** | 4 | 4 |  |
| **Age (median [IQR])** | 37.11 [32.00, 41.76] | 28.63 [23.50, 34.83] | 0.386 |
| **Male (%)** | 4 (100.0) | 3 ( 75.0) | 1.000 |
| **Prior Therapies (median [IQR])** | 4.50 [3.75, 5.50] | 5.00 [4.75, 5.00] | 0.757 |
| **AutoSCT(%)** | 4 (100.0) | 2 ( 50.0) | 0.414 |
| **Radiotherapy (%)** | 1 ( 25.0) | 1 ( 25.0) | 1.000 |
| **BV (%)** | 4 (100.0) | 4 (100.0) | NA |
| **BV+auto (%)** | 4 (100.0) | 2 ( 50.0) | 0.414 |
| **Advanced stage (%)** | 3 ( 75.0) | 4 (100.0) | 1.000 |
| **Refractory to first line (%)** | 2 ( 50.0) | 3 ( 75.0) | 1.000 |
| **Refractory to last line (%)** | 2 ( 50.0) | 2 ( 50.0) | 1.000 |
| **Extranodal (%)** | 2 ( 50.0) | 3 ( 75.0) | 1.000 |
| **B-Symptoms (%)** | 1 ( 25.0) | 1 ( 25.0) | 1.000 |

## References

1. Weniger, M. A. & Küppers, R. Molecular biology of Hodgkin lymphoma. Leukemia 35, 968– 981 (2021).

2. Alig, S. K. et al. Distinct Hodgkin lymphoma subtypes defined by noninvasive genomic profiling. Nature 625, 778–787 (2024).

3. Menéndez, V., Solórzano, J. L., Fernández, S., Montalbán, C. & García, J. F. The Hodgkin Lymphoma Immune Microenvironment: Turning Bad News into Good. Cancers 14, 1360 (2022).

4. Aoki, T. & Steidl, C. Novel insights into Hodgkin lymphoma biology by single-cell analysis. Blood 141, 1791–1801 (2023).

5. Cha, J.-H., Chan, L.-C., Li, C.-W., Hsu, J. L. & Hung, M.-C. Mechanisms Controlling PD-L1 Expression in Cancer. Molecular Cell 76, 359–370 (2019).

6. Opinto, G. et al. Hodgkin Lymphoma: A Special Microenvironment. JCM 10, 4665 (2021).

7. Ma, Y. et al. The CD4+CD26− T-cell population in classical Hodgkin’s lymphoma displays a distinctive regulatory T-cell profile. Laboratory Investigation 88, 482–490 (2008).

8. Veldman, J. et al. CD4+ T cells in classical Hodgkin lymphoma express exhaustion associated transcription factors TOX and TOX2: Characterizing CD4+ T cells in Hodgkin lymphoma. OncoImmunology 11, 2033433 (2022).

9. Zuazo, M. et al. Systemic CD4 Immunity as a Key Contributor to PD-L1/PD-1 Blockade Immunotherapy Efficacy. Front. Immunol. 11, 586907 (2020).

10. Maharaj, K., Uriepero, A., Sahakian, E. & Pinilla-Ibarz, J. Regulatory T cells (Tregs) in lymphoid malignancies and the impact of novel therapies. Front. Immunol. 13, 943354 (2022).

11. Cader, F. Z. et al. Mass cytometry of Hodgkin lymphoma reveals a CD4+ regulatory T-cell–rich and exhausted T-effector microenvironment. Blood 132, 825–836 (2018).

12. Topalian, S. L. et al. Safety, Activity, and Immune Correlates of Anti–PD-1 Antibody in Cancer. N Engl J Med 366, 2443–2454 (2012).

13. Brahmer, J. R. et al. Safety and Activity of Anti–PD-L1 Antibody in Patients with Advanced Cancer. N Engl J Med 366, 2455–2465 (2012).

14. Ansell, S. M. et al. PD-1 Blockade with Nivolumab in Relapsed or Refractory Hodgkin’s Lymphoma. N Engl J Med 372, 311–319 (2015).

15. Armand, P. et al. Programmed Death-1 Blockade With Pembrolizumab in Patients With Classical Hodgkin Lymphoma After Brentuximab Vedotin Failure. JCO 34, 3733–3739 (2016).

16. Xu-Monette, Z. Y., Zhang, M., Li, J. & Young, K. H. PD-1/PD-L1 Blockade: Have We Found the Key to Unleash the Antitumor Immune Response? Front. Immunol. 8, 1597 (2017).

17. Bryan, L. J. et al. Pembrolizumab Added to Ifosfamide, Carboplatin, and Etoposide Chemotherapy for Relapsed or Refractory Classic Hodgkin Lymphoma: A Multi-institutional Phase 2 Investigator-Initiated Nonrandomized Clinical Trial. JAMA Oncol 9, 683 (2023).

18. Calabretta, E. et al. Chemotherapy after PD -1 inhibitors in relapsed/refractory Hodgkin lymphoma: Outcomes and clonal evolution dynamics. Br J Haematol 198, 82–92 (2022).

19. Al-Hadidi, S. A. & Lee, H. J. Checkpoint Inhibition Therapy in Transplant-Ineligible Relapsed or Refractory Classic Hodgkin Lymphoma. JCO Oncology Practice 17, 64–71 (2021).

20. Milrod, C. J., Pelcovits, A. & Ollila, T. A. Immune checkpoint inhibitors in advanced and relapsed/refractory Hodgkin lymphoma: current applications and future prospects. Front. Oncol. 14, 1397053 (2024).

21. Hwang, B., Lee, J. H. & Bang, D. Single-cell RNA sequencing technologies and bioinformatics pipelines. Exp Mol Med 50, 1–14 (2018).

22. Tu, A. A. et al. TCR sequencing paired with massively parallel 3′ RNA-seq reveals clonotypic T cell signatures. Nat Immunol 20, 1692–1699 (2019).

23. Armand, P. et al. Nivolumab for Relapsed/Refractory Classic Hodgkin Lymphoma After Failure of Autologous Hematopoietic Cell Transplantation: Extended Follow-Up of the Multicohort Single-Arm Phase II CheckMate 205 Trial. JCO 36, 1428–1439 (2018).

24. Cheson, B. D. et al. Recommendations for Initial Evaluation, Staging, and Response Assessment of Hodgkin and Non-Hodgkin Lymphoma: The Lugano Classification. JCO 32, 3059–3067 (2014).

25. World Medical Association Declaration of Helsinki: Ethical Principles for Medical Research Involving Human Subjects. JAMA 310, 2191 (2013).

26. Robert, C. et al. Nivolumab in Previously Untreated Melanoma without *BRAF* Mutation. N Engl J Med 372, 320–330 (2015).

27. Satija, R., Farrell, J. A., Gennert, D., Schier, A. F. & Regev, A. Spatial reconstruction of single-cell gene expression data. Nat Biotechnol 33, 495–502 (2015).

28. Sturm, G. et al. Scirpy: a Scanpy extension for analyzing single-cell T-cell receptor-sequencing data. Bioinformatics 36, 4817–4818 (2020).

29. Younes, A. et al. Nivolumab for classical Hodgkin’s lymphoma after failure of both autologous stem-cell transplantation and brentuximab vedotin: a multicentre, multicohort, single-arm phase 2 trial. The Lancet Oncology 17, 1283–1294 (2016).

30. Fisler, D., Sikaria, D., Yavorski, J., Tu, Y. & Blanck, G. Elucidating feed-forward apoptosis signatures in breast cancer datasets: Higher FOS expression associated with a better outcome. Oncol Lett (2018) doi:10.3892/ol.2018.8957.

31. Joshi, N. S. et al. Inflammation Directs Memory Precursor and Short-Lived Effector CD8+ T Cell Fates via the Graded Expression of T-bet Transcription Factor. Immunity 27, 281–295 (2007).

32. Renkema, K. R. et al. KLRG1+ Memory CD8 T Cells Combine Properties of Short-Lived Effectors and Long-Lived Memory. The Journal of Immunology 205, 1059–1069 (2020).

33. Papavassiliou, A. G. & Musti, A. M. The Multifaceted Output of c-Jun Biological Activity: Focus at the Junction of CD8 T Cell Activation and Exhaustion. Cells 9, 2470 (2020).

34. Shresta, S., Graubert, T. A., Thomas, D. A., Raptis, S. Z. & Ley, T. J. Granzyme A Initiates an Alternative Pathway for Granule-Mediated Apoptosis. Immunity 10, 595–605 (1999).

35. Zhang, Z. et al. T Cell Dysfunction and Exhaustion in Cancer. Front. Cell Dev. Biol. 8, 17 (2020).

36. Valpione, S. et al. Immune awakening revealed by peripheral T cell dynamics after one cycle of immunotherapy. Nat Cancer 1, 210–221 (2020).

